# Prescribing pre- and post-operative physical activity interventions for people undergoing breast cancer surgery: a qualitative systematic review

**DOI:** 10.1101/2023.05.16.23290041

**Authors:** Lauren Howe, Andy Husband, Anna Robinson-Barella

**Author notes:** **Author emails**: Lauren Howe Andy Husband Anna Robinson-Barella (corresponding author). corresponding author Mrs Anna Robinson-Barella (corresponding author) ORCID iD: 0000-0002-9523-4760. Full postal address: School of Pharmacy, King George VI Building, Newcastle University, Newcastle upon Tyne, NE1 7RU, UK. **Data availability statement:** The data that support the findings of this study are available from the corresponding author upon reasonable request. **Funding statement:** This work was undertaken as part of a Masters of Pharmacy Undergraduate degree (author LH, supervised by author AR-B); there was no funding for this work. **Conflict of interest disclosure:** None. **Ethics approval statement:** N/A. **Patient consent statement:** This work is a qualitative systematic review, thus there was no direct patient involvement in this work. **Permission to reproduce material from other sources:** Not applicable; this work is original. **Clinical trial registration:** Not applicable. **Author contributions:** LH led on the day-to-day running of the project, data collection and writing of this manuscript. AR-B oversaw the running of this project as Principal Investigator and provided project management expertise. AR-B and AH provided methodological input and expertise. All authors read, provided comments on, and approved the final manuscript.

## Abstract

**Introduction:** Undertaking physical activity, pre- and post-operatively, can benefit recovery time and improve post-surgical outcomes. One cohort of patients that have reported these benefits are those undergoing surgery for breast cancer. Yet, what remains unclear is the level to which physical activity interventions are implemented into standard surgical care for patients with breast cancer.

**Aims:** This systematic review aimed to examine existing qualitative evidence focusing on pre- and post-operative physical activity interventions to better understand the benefits and shortcomings of physical activity within the surgical journey.

**Methods:** A systematic literature search was undertaken in November 2022, across five databases: MEDLINE, PsycINFO, Embase, CINAHL and Scopus. Qualitative studies involving people with breast cancer who had undertaken a physical activity intervention, either pre- and/or post-operatively, were included for analysis. The review was registered on PROSPERO: CRD42022372466 and performed according to PRISMA guidelines. The Critical Appraisal Skills Programme qualitative study checklist was used to assess study quality.

**Results:** Fourteen studies were included, comprising the perspectives of 418 people receiving surgery for breast cancer. One study implemented pre-operative physical activity interventions; the remaining studies focused on post-operative interventions. A narrative systematic review was undertaken due to heterogeneity in reported results. Four themes were developed by thematic analysis, centring on: (i) factors promoting engagement with physical activity interventions; (ii) factors preventing engagement with physical activity interventions; (iii) the impact of pre- and post-operative interventions on physical and psychological health; and (iii) participant recommendations for pre- and post-operative interventions.

**Conclusion:** pre- and post-operative physical activity interventions were well-accepted. Patients recognised factors which promoted or prevented engagement with interventions, as well as pre- and post-operative physical and psychological benefits that arose as a result. Evidence based co-design studies may further inform successful implementation of physical activity into standard care for surgical breast cancer patients.

## Introduction

It is estimated that there are on average 375,400 new cancer cases each year in the United Kingdom (UK), with 55,920 of these newly diagnosed cases being breast cancer.^1^ In 2017, breast cancer counted for 15.1% of all new malignant cancer registrations (compared to 13.5% of registrations of prostate cancer, 12.7% lung cancer, 5.9% gynaecological cancers) and it has been the most frequently diagnosed cancer in England since it overtook lung cancer in 1996.^2^ Similar statistics can be seen on a worldwide scale with an estimated 2.3 million new breast cancer cases each year.^3^ The National Health Service (NHS) Long Term Plan has placed emphasis on new guidelines for cancer care, given the growing incidence of people in the UK being diagnosed with cancer each year.^4^ As well as emphasising the importance of early cancer diagnosis to improve rates of cancer survival,^4^ the Long Term Plan hopes to provide every person diagnosed with cancer personalised care; reference to this included a health and wellbeing care plan, alongside follow-up treatment after surgery.^4^

Pre-operative and post-operative surgical care for breast cancer patients is complex and challenging, but is important in improving recovery and reducing morbidity.^5^ The Enhanced Recovery After Surgery (ERAS) society undertook a systematic review in 2017 in order to present evidence-based recommendations for the pre- and post-operative management of breast cancer surgery.^6^ Their key recommendations included: use of opioid-sparing medication, minimal preoperative fasting, use of anaesthetic techniques that decrease nausea and vomiting, early mobilization post-surgery and post-discharge home support with early physical activity.^6^ The prescription of physical activity in healthcare settings is an area growing in research over recent years, it has been shown to reduce the incidence of chronic disease development and improve quality of life.^7, 8^

Physical activity to support patients undergoing breast cancer surgery has been proven beneficial. Findings from current literature have seen patients report reduced pain and improved quality of life post-surgery,^9,10^ as well as improvements in their physical functions like strength, stamina, and cardiovascular fitness, and a reduction in cancer-related fatigue.^11–13^ Yet, despite the evidence that demonstrates the benefits arising from pre- and post-operative physical activity, challenges remain that affect patient adherence and engagement with physical activity interventions.^14^ Teo *et al*. found that patient motivation and capability were barriers for patient engagement with prescribed exercise programmes.^15^ Another reported barrier which affects the uptake of physical activity interventions is the willingness of healthcare professional to recommend physical activity; one UK study from 2008 reported that approximately half of oncologists and surgeons do not routinely discuss physical activity with their patients.^16^ This led to independent parties and charities, such as Macmillan Cancer Support, publishing evidence-based exercise advice and guidance for patients. Specifically, this focused on the integration of pre- and post-operative physical activity in the cancer care pathway.^17^

This systematic review takes the form of a qualitative narrative synthesis examining physical activity interventions to support patients undergoing breast cancer surgery. The focus will be on both the pre-operative and post-operative time frame to understand the full scope of breast cancer care throughout the surgical pathway. This work seeks to assist in informing future clinical decisions on the implementation of physical activity interventions into standard care for breast surgery patients, and ultimately improve the quality of care that patients receive.

## Method

### Protocol registration

This systematic review is registered with PROSPERO (Registration number CRD42022372466) and has been conducted in accordance with the ‘Preferred Reporting Items for Systematic Reviews and Meta-Analyses (PRISMA)’ guidelines (see Appendix A in the Supplementary File).^18^

### Eligibility criteria

To inform the eligibility criteria, PICOS (Population, Intervention, Comparator, Outcome, Study type) criteria were developed (Table 1). Qualitative studies with a population of patients with breast cancer who were due to undergo surgery, or had already undergone surgery, were included. There were no limits placed on patient’s age, sex, ethnicity or country of surgery. As outlined in Table 1, any type of breast cancer surgery was acceptable.

**Table 1:** Eligibility of studies: PICOS criteria.

|  |  |
| --- | --- |
| **Population** | • Men or Women with who have undergone or are scheduled to have breast cancer surgery.<br>• All surgery types included: mastectomy (total or partial), breast conserving surgery, sentinel lymph node biopsy, anterior lymph node dissection and combinations. |
| **Intervention** | • Any form of physical activity undertaken in the peri-operative period (from moment surgery is contemplated up to full recovery).<br>• Physical activity is defined by the World Health Organisation as any bodily movement that requires energy expenditure.<sup>60</sup> Consequently, any intervention that meets this definition was included. Examples identified include running, swimming, walking, stretching and resistance training.<br>• Interventions could have taken place up to 5 years post-surgery as this is the length of time cancer patients remain under care of oncology team according to NICE guidelines.<sup>19</sup> |
| **Comparator** | • Control arm was not necessary – purposely kept open for ethical reasons and current practice of care.<br>• Comparison could be alternative physical activity types or ‘usual care’. |
| **Outcome** | • Any reported qualitative outcome from patients including quality of life, fatigue, mental wellbeing, impact on recovery. |
| **Study Type** | • Qualitative methodologies including but not limited to: focus groups, semi-structured interviews.<br>• All quantitative methodologies will be excluded, this includes randomised controlled trials.<br>• Systematic reviews will also be excluded. |

The studies must have demonstrated a form of physical activity intervention within the surgical pathway, either pre-or post-operatively. There were no limits placed on the type of physical activity or the delivery method of the intervention. Studies were included if the physical activity took place up to 5 years post-surgery; this decision was based on NICE guidelines as a typical post-operative time frame where patients would remain under the care of the oncology team.^19^ The authors recognised that some studies with a longer time frame were still eligible for inclusion in this review, given that these participants were reported to still be in recovery from the surgery required as part of breast cancer treatment. Included studies were not required to have a control or comparator group; ‘usual care’ or an alternative intervention was deemed an accepted comparator.

Only qualitative studies were eligible for inclusion; quantitative studies, included but not limited to, randomised controlled trials, observational studies, longitudinal studies, and prospective cohort studies were excluded. Mixed-methods studies were included if the qualitative data was sufficient. Studies that were not available in the English Language were excluded.

### Search strategy, information sources and study selection

A systematic literature search was conducted in November 2022 (by author LH) across five electronic databases: MEDLINE, PsycINFO, CINAHL, Embase and Scopus (searches were conducted from journal conception to present day). Additional papers were identified via the grey literature within reference lists and personal libraries of authors.

The search terms were created by the authors, with the support of a medical librarian. The search terms were created in the English Language using Medical Subject (MeSH) headings and keywords, with the search strategy compiled using Boolean operators (AND/OR). Example the search strategy and journal-specific search terms are included in Appendix B of the Supplementary File. All records were converted into the reference database EndNote (Version X9) for ease of management.

Title and abstracts of all papers obtained were reviewed by one author (LH) to assess eligibility. Full texts were retrieved for articles that met the inclusion criteria for further evaluation, and for those that could not be rejected with certainty. Full text articles were screened independently by two authors (LH and AR-B); disagreements were resolved by discussion with a third author (AH) where necessary.

### Data extraction

Data extraction was undertaken by study authors using a customised data extraction form, amended from a previous review in this field (Appendix C in the Supplementary File).^20^ Data extracted included: year of publication, population, participant characteristics, type of exercise intervention, details of exercise intervention, pre- and/or post-operative intervention point, details of additional interventions and control group. Study interventions were grouped into one of three possible timepoints within the surgical pathway for analysis: pre-operative intervention timing (implemented before the surgical procedure), post-operative timing (implemented after the surgical procedure), or a combination of both pre- and post-operative (implemented before and continued after operation).

### Quality appraisal

The included studies were appraised for methodological quality using the Critical Appraisal Skills Programme (CASP) qualitative study checklist.^21^ Any uncertainties were discussed between authors (LH and AR-B). All studies were assigned with a methodological quality score for ease of reporting, expressed as a percentage.

### Data analysis and synthesis

Authors sought to establish further understanding on the implementation of physical activity interventions within surgical care pathways. This review takes the form of a narrative synthesis, underpinned by thematic analysis, where authors developed, defined and refined key themes within the existing qualitative data. Heterogeneous qualitative measures were reported within the included studies, so a meta-analysis was not possible.

## Results

### Search results

The initial search obtained 1,114 records from 5 databases: Medline, PsycINFO, CINAHL, EMBASE and Scopus. An additional 35 records were obtained from grey literature. After removal of duplicates, 965 records remained, of which 756 were inappropriate and thus excluded from the review. Assessment of the remaining 208 full text articles gave 14 records eligible for review which fit the criteria. A PRISMA flowchart (Figure 1) was created to illustrate the search process from start to finish, with reasons for exclusion included at each step.

**Figure 1:**
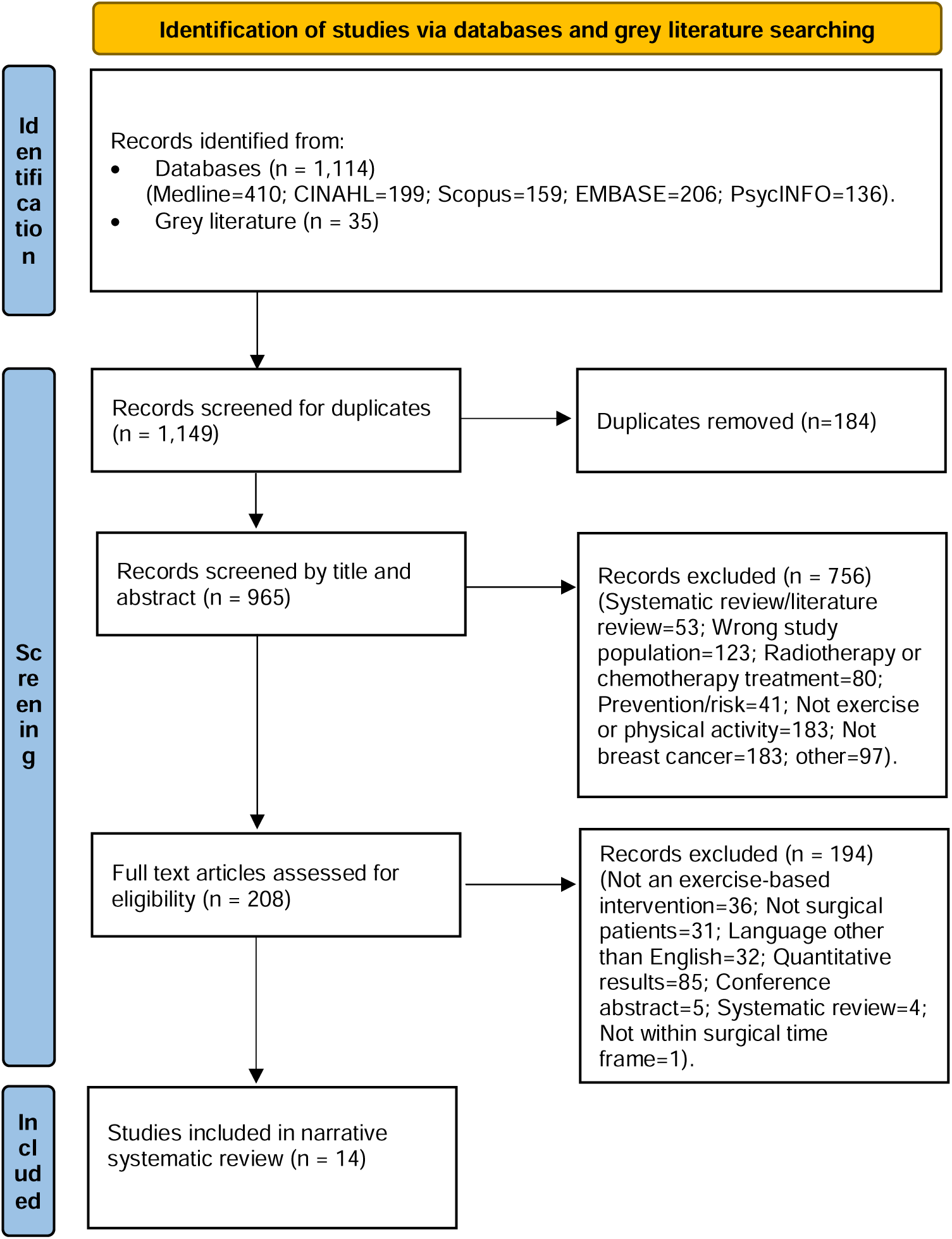
PRISMA Flowchart.

### Study characteristics

In total, 14 studies were included within this systematic review (see Table 2 for Study Characteristics). The studies spanned 7 different countries, including Canada,^22–24^ USA,^25–28^ Sweden,^29, 30^ Ireland,^31^ England,^32^ Korea^33, 34^ and Scotland,^35^ and were conducted between the years of 2008 to 2021. The included studies shared the perspectives of 418 breast cancer patients who had either undergone, or were about to undergo, surgery as a treatment for breast cancer. The reported age range of the participants was 25-82 years old. Of the included studies, only one explored and implemented a pre-operative physical activity intervention;^22^ the remaining thirteen studies focused on interventions that occurred post-operatively. Across the included papers, 5 different terminologies were used to describe the type of breast cancer surgery; these included mastectomy (or total mastectomy);^22, 25–28, 30–34^ partial mastectomy (or breast conserving surgery or lumpectomy);^22, 25–28, 30–34^ reconstruction;^26–28, 31^ sentinel lymph node biopsy;^22, 28, 29, 32, 33^ and axillary node dissection/clearance.^22, 28, 29, 32, 33^ Three of the papers did not specify a surgery type.^23, 24, 35^

**Table 2:**
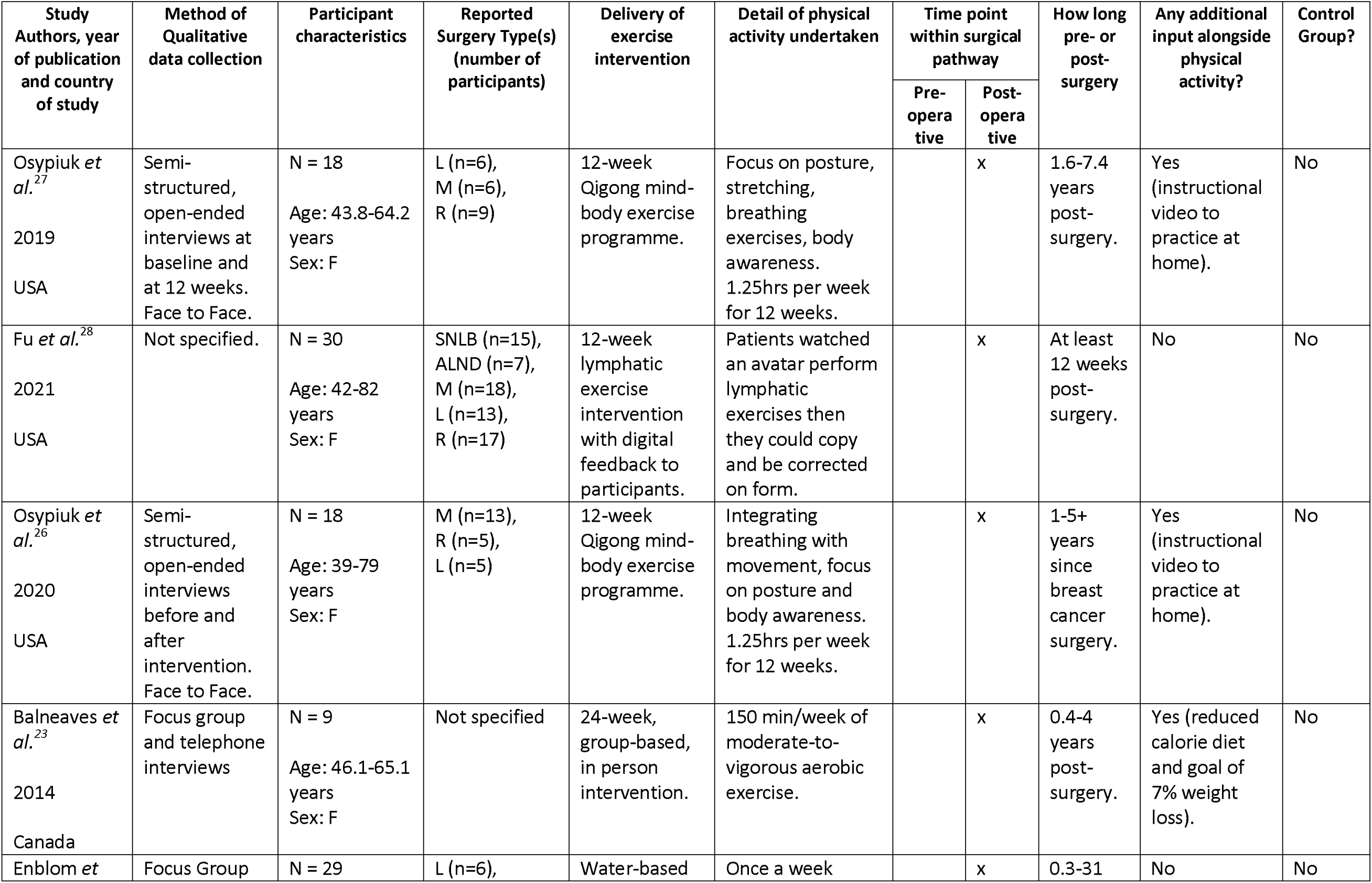
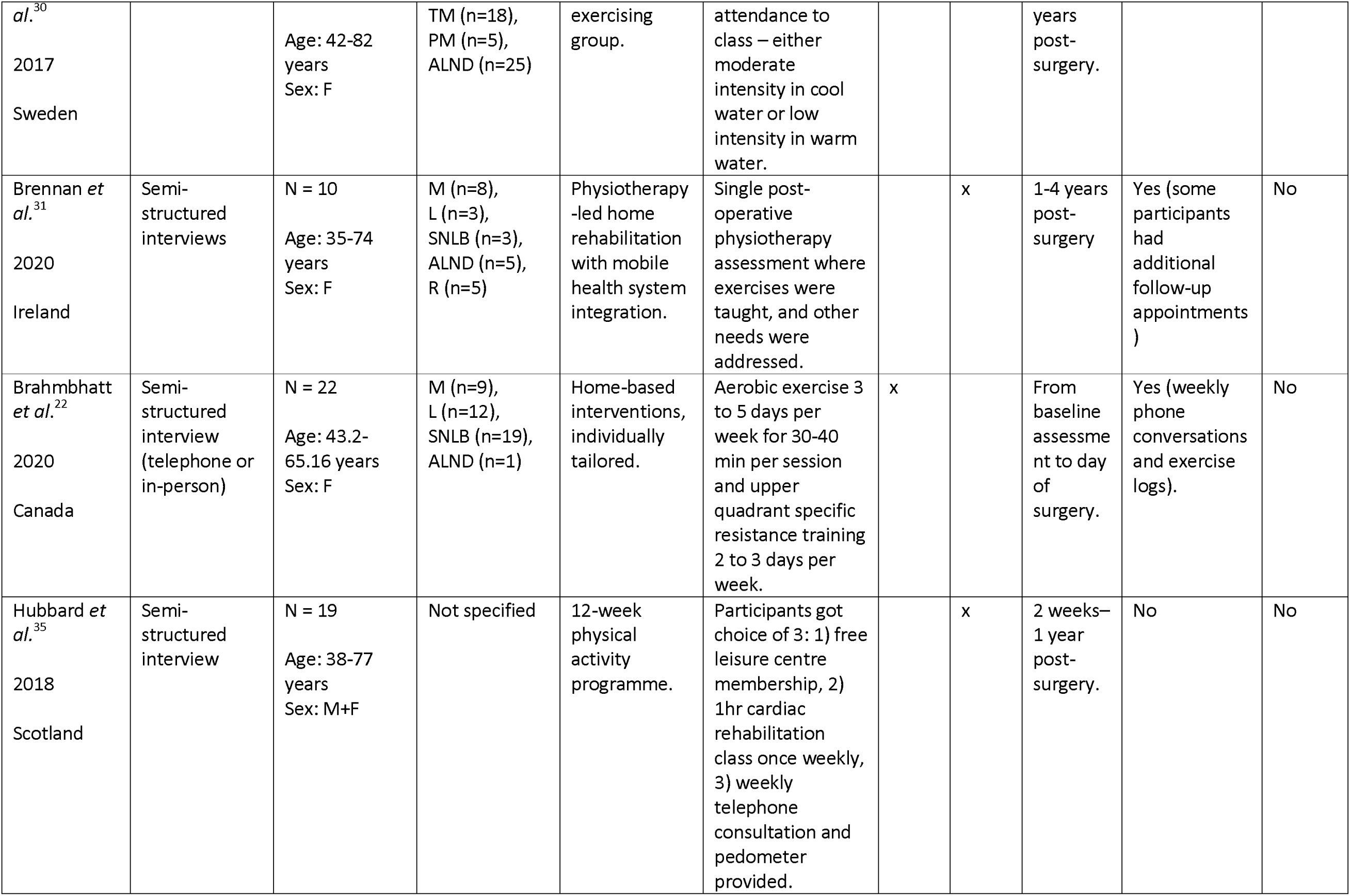
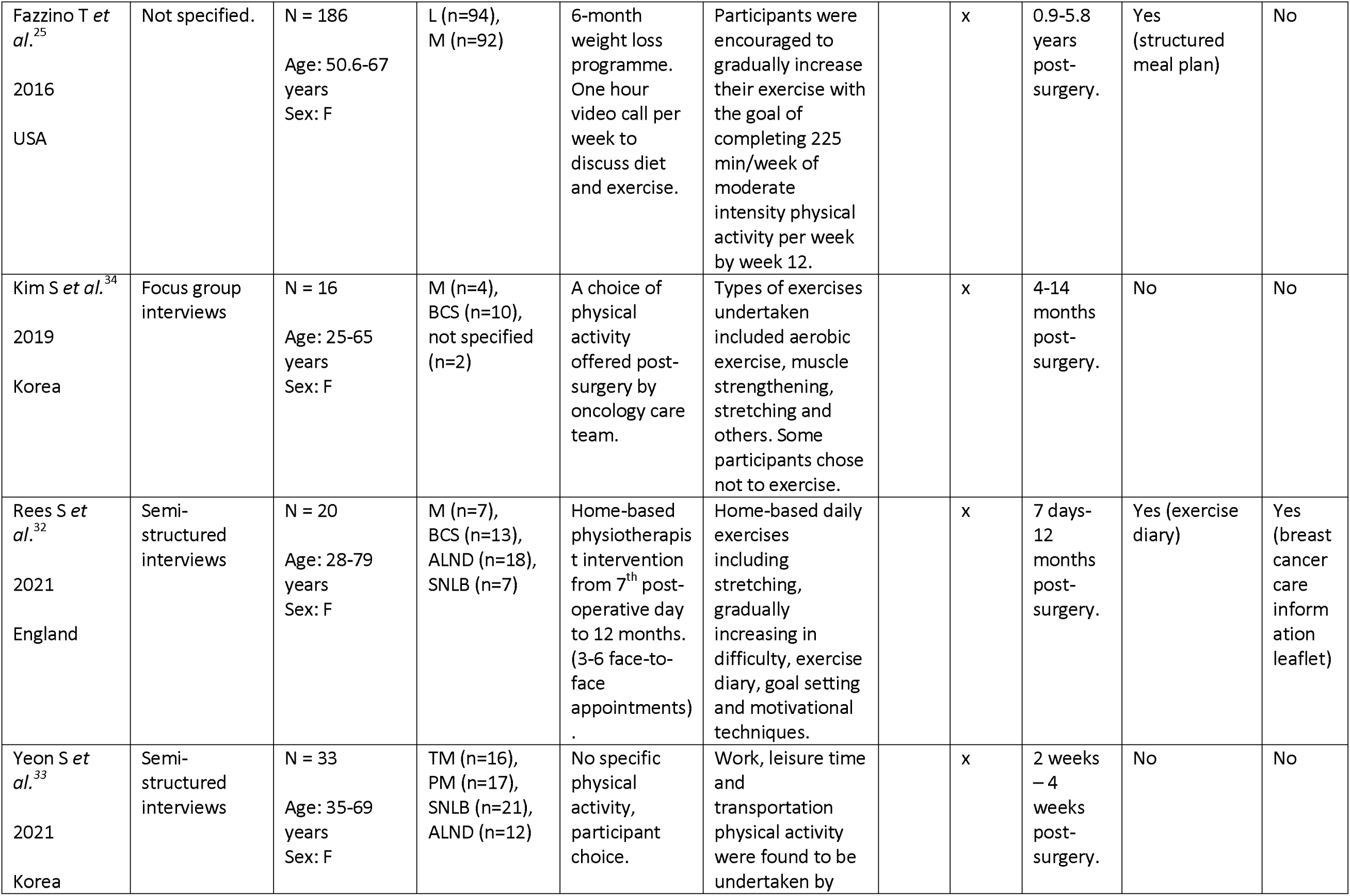
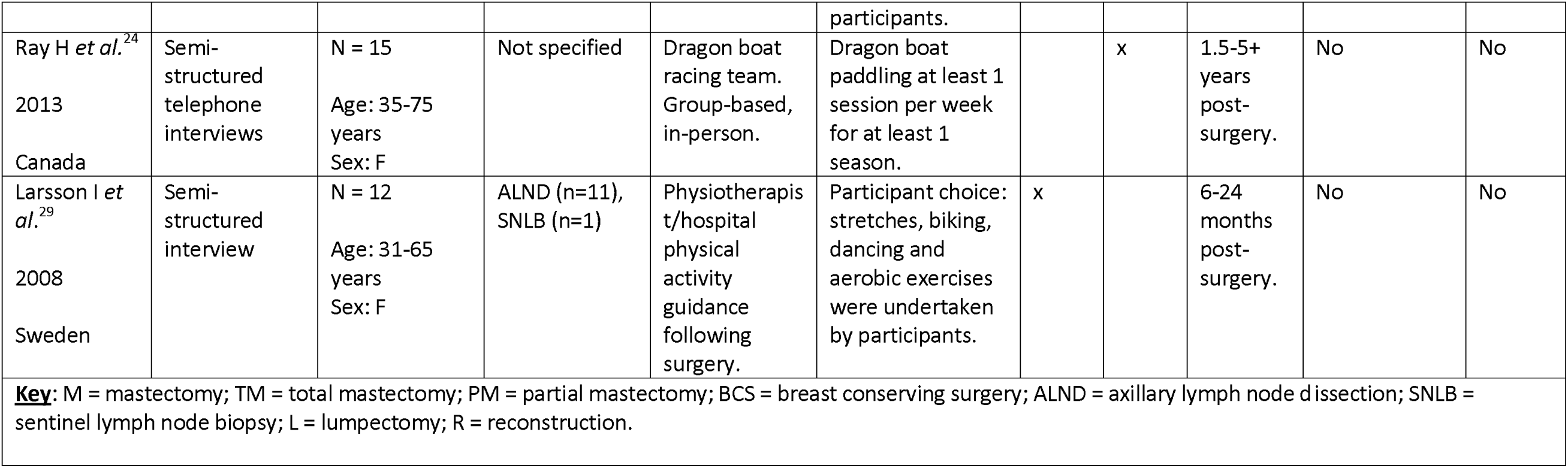
Study characteristics (adapted from a surgical data extraction form featured in a systematic review by the research team).^46^.

With regards to the physical activity intervention explored, there was variation in both the (i) type and (ii) delivery method used; two studies undertook vigorous aerobic exercise as their intervention;^22, 23^ one study focused on water-based exercise classes;^30^ two of the articles studied physiotherapist-led exercises;^32^ and one of these studies also implemented digital strategies via a mobile health intervention.^31^ Other interventions with digital elements included kinetic lymphatic exercises,^28^ and a weight loss programme over 6 months with weekly video call sessions.^25^ Some of the studies opted for low-intensity physical activity, exploring mind-body exercising (breathing and stretching focused exercise)^26, 27^ and group dragon boat racing.^24^ Three studies did not specify a particular physical activity as part of their intervention; instead, participants were given the opportunity to either choose from 3 presented intervention choices^35^ or undertake any physical activity they desired after post-surgical advice was provided.^33, 34^ Delivery of the interventions also showed variation, with five studies using face-to-face classes,^23, 24, 26, 27, 30^ five using remote delivery methods^22, 25, 28, 31, 32^ and four interventions included a combination of in-person and home-based elements.^29, 33–35^

There was also a range of qualitative methodologies used within the studies, including semi-structured interviews^22, 24, 26, 27, 29, 31–33, 35^ and focus groups,^23, 30, 34^ with 2 studies not specifying their methodological approach.^25, 28^ Only one of the studies had a control group comparator.^32^

### Study quality

The methodological quality of the 14 included studies was deemed to be ‘good’, with a mean Critical Appraisal score of 75% (see Appendix D in the Supplementary File). Quality assessment scores across the included papers ranged from 40%^25^ to 100%.^33^ The poorest performing area amongst the studies was the analysis of the relationship between researcher and participants; only 8 of the 14 papers considered this.^23, 24, 27, 28, 30, 32–34^ The highest performing areas were data analysis, data collection and use of qualitative methodology.

### Narrative synthesis

Four overarching themes were developed from thematic analysis of the data, which centred on: (i) factors promoting patient engagement with physical activity interventions; (ii) factors preventing patient engagement with physical activity interventions; (iii) the impact of pre- and post-operative interventions on a person’s physical and psychological health; and (iv) patient recommendations for pre- and post-operative physical activity interventions. Each theme will be discussed in turn, with participant quotes integrated within the text to ensure that the patient voice remained at the forefront of the results reported (see Figure 2).

**Figure 2:**
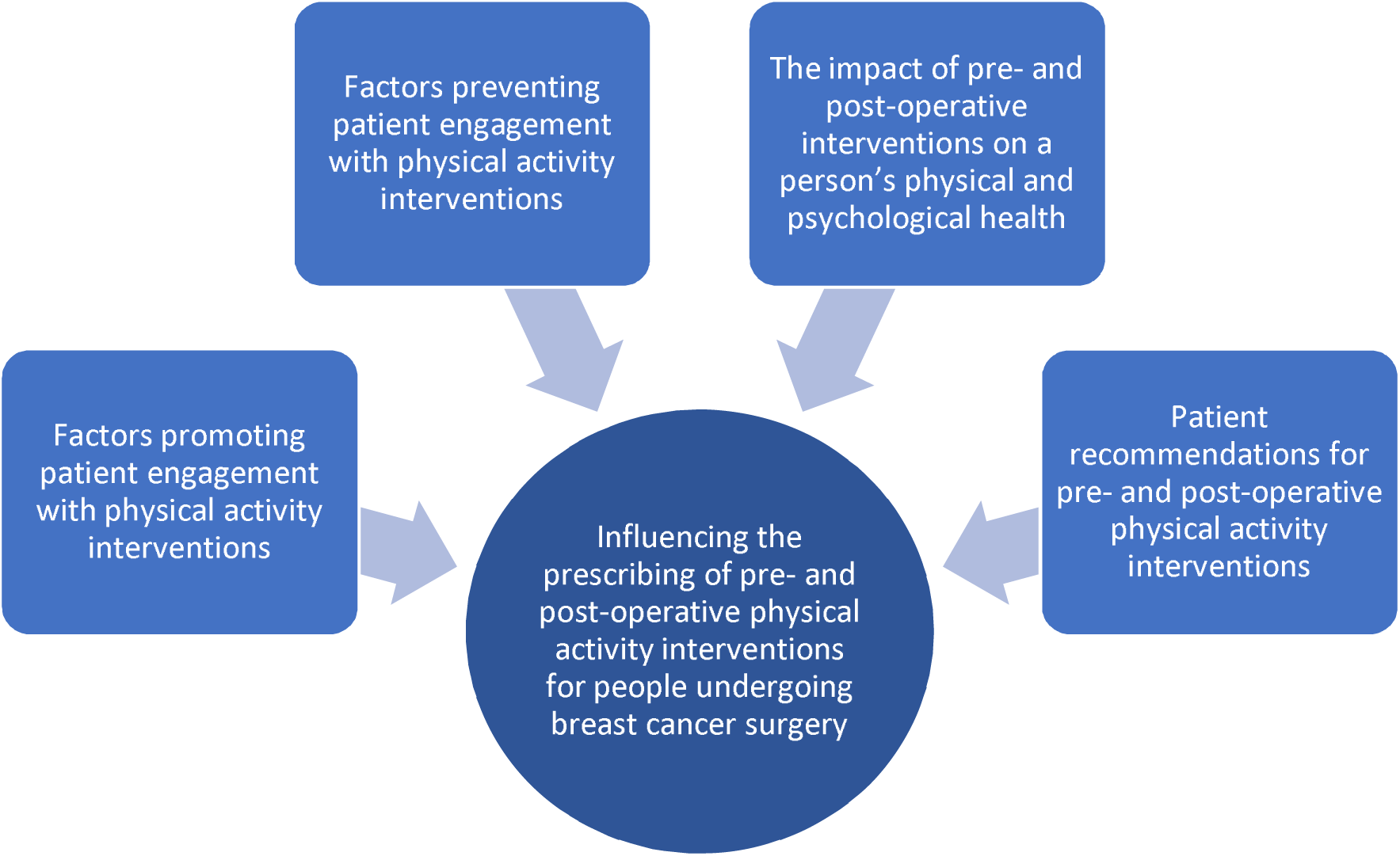
Four overarching themes influencing the prescribing of pre- and post-operative physical activity interventions for patients undergoing breast cancer surgery.

### Theme 1: Factors promoting patient engagement with physical activity interventions

All 14 papers reported factors which made the participants more likely to attend, participate, and complete the physical activity intervention. Four of the papers reported increased engagement rates if the exercises were easy to follow and achievable.^22, 26–28^ One participant described having the ability to modify and adapt the exercises so that *“anyone could have done (the exercises), which made it really great for any woman, of any age, at any physical level”*.^22^ Another commented how the sense of achievement of progressing onto more challenging exercises facilitated further adherence.^32^ One paper postulated that easier exercises improved participant self-satisfaction and rates of adherence, as they did not experience physical limitations – something that patients with breast cancer reportedly struggled with post-operatively. ^26^

> “It’s acknowledging [your body] more and … all the amazing things it’s doing (exercises), and not focusing on what it’s not doing or what you can’t do”.^26^

An individual’s internal motivation and desire to recover, alongside fear of cancer recurrence, facilitated adherence to physical activity interventions in almost half of the papers.^23, 25, 29, 31, 33, 34^ Patients expressed needing to know they had *“done everything they could”* to prevent cancer recurrence;^23^ viewing exercise as a perceived solution to this, increased participant rates.^32–34^

> “it was like looking down the barrel of a gun and you’ve got your thumb on the trigger. Are you gonna pull it and just eat whatever you want and do whatever you were doing previously? Or am I gonna really take a hold of what I’m doing and do whatever I could? So, for me, it was fear. I’m in control of this and I can go this way or I can follow a healthy lifestyle.” ^23^

Emphasis was placed on the trust participants had in an intervention if it had come from the hospital care team, with oncology trained professionals and breast cancer specific exercises.^22, 31, 32, 34^ If the physical activity was pitched as a compulsory part of their cancer treatment, rather than a choice, participants expressed this made them more likely to participate; *“if you say to me this is what you need to do to get better, I’ll do it”.*^31^ Participants perceiving the completion of physical activity as a non-cancer related activity, was also reported as a factor which encouraged participation in physical activity. ^23, 29^

Group-based physical activity interventions were popular with participants and were associated with facilitating attendance and participation across 9 of the 14 papers.^23–26, 30, 31, 33–35^ The supportive and relatable interaction with other patients undergoing surgery for breast cancer was deemed most useful, with one participant describing *“meeting people who have all been through similar, you can all relate”.*^31^ Another reported comparing themselves to other participants in the group as a means of feeling re-assurance, as *“I could see that I was actually doing this (exercise) too, and I could keep up with everybody else”*.^28^ While most papers referred to the interaction with other patients undergoing breast cancer surgery as advantageous, in the study by Hubbard *et al.,* one female participant described preferring to exercise with the general public in a leisure centre; it gave her a break from being surrounded by cancer.^35^

Friends and family were also mentioned by three of the papers as a form of extended support network which promoted a person’s participation in physical activity;^23, 33, 35^ the inclusion of family members provided support at home, in addition to that when attending their in-person intervention.^23^

> “You can get [slight laugh] very competitive with yourself, and especially with my, my father-in-law … he’s got an App on his phone, so he started just, you know: “oh, I’ve done this many steps,” I’d be like “well if he’s done that many steps, I’ve got to do …,” so you do … it’s benefitted everybody”.^35^

Additional support measures from intervention leaders were found to facilitate improved adherence; for example, interventions that utilised check-ins, continued feedback and monitoring were perceived as beneficial.^22, 23, 28, 33^ Constant re-assurance was a key factor with patients reporting they *“like the fact it tells you if you are doing the exercises correctly”*.^28^ This gentle guidance from intervention providers was shown to enhance participant confidence in their own ability to complete the exercises. Further factors identified as facilitators of adherence to interventions are detailed in Appendix E of the Supplementary File.

### Theme 2: Factors preventing patient engagement with physical activity interventions

Twelve of the 14 included studies commented on factors that hindered adherence to the prescribed physical activity interventions. One recurring theme that related to pain, fatigue and other cancer treatment-related side effects was mentioned by participants in 9 of the studies.^23, 25, 27, 29, 31–35^ Physical limitations experienced post-operatively, including pain at the surgical site, caused adherence issues in many patients;^26, 27, 29, 31–33^ consequently, participants struggled to complete the exercises as *“if I did anything strenuous, it hurt and I felt tightness here… it was frustrating”*.^29^ Cancer-related fatigue was also reported by half of the papers as a key factor preventing physical activity in patients with breast cancer.^23, 25, 29, 31, 32, 34, 35^ The subsequent exhaustion and compromised fitness level meant they were unable to engage with their prescribed physical activity with one female participant reporting *“some days, I feel like I’d faint…When really exhausted, you have to sleep… it’s just like playing dead”.*^34^ It was also reported in one paper that the fatigue then made the patients feel like *“an outsider”* when exercising with the public because they could not complete the exercise as efficiently as others.^34^

Due to its adverse effects making the pre- and post-operative physical activity more strenuous, chemotherapy adjuvant treatment was reported as a barrier to completion of the physical activity.^29, 31, 32^ Participants reported the physical activity to be more manageable post-surgery, until the point in their treatment that chemotherapy began; *“the last three weeks with the first lot of chemo this…[doing the exercises has] been a lot harder than I ever anticipated”*.^32^ In addition, the mental concentration, required for the physical activity intervention was perceived as a challenge due to *“chemo brain”*, a term one patient used to describe her diminished cognitive function due to the chemotherapy treatment.^23^

Fears and myths surrounding exercise whilst undergoing surgical treatment for breast cancer were widely recognised,^23, 29, 31, 32, 34^ with inadequate information on the safety of post-operative exercise being a main contributor. In particular, participants in two papers spoke of fear that the exercise would hinder their recovery.^33, 34^ One paper reported kinesiophobia (a fear of movement) that women experienced in the acute period following surgical treatment for breast cancer, impeding their participation in physical activity.^32^ Despite the prevention of cancer recurrence through exercise being a previously reported motivator to participation in the physical activity interventions, fear of cancer recurrence, as a result of exercising, was also a reported barrier to participation in physical activity. Some patients reported that exercising could cause disease recurrence; “I had a friend who had recurrence and she believed it was because she exercised too aggressively.”^34^ In this same paper, patients also believed carrying heavy things could cause lymphoedema. Inadequate information and education caused fears and myths to circulate around cancer patients and caused lack of confidence in completion of the prescribed exercises; *“I didn’t even know whether I was doing it right or not, you know sitting on the bed supposed to be doing this or this, or whatever, I didn’t bother then”*.^31^

> “if I exercise too much, the energy I’m supposed to use to beat the cancer, if it gets used up for exercise, that’s probably worse for my body…I’ve talked with a lot of patients, and heard that if you exercise too much, you can get recurrence”.^34^

Fear reported by family members was also recognised as a barrier preventing patients adhering to physical activity interventions. One participant described how family associated visual changes as a result of exercise (including weight loss) with disease recurrence, asking *“you’re losing weight, oh no, you’re losing weight! Are you okay?”.*^23^

Recurring barriers within studies included patients feeling self-conscious regarding their body-image as a result of mastectomy, hair loss, or surgical scars.^33–35^ In particular, patients spoke of the discomfort and *“hindrance”* of exercising in a wig.^34^ In another paper, a patient commented how the presence of a drain attached to their body due to post-surgical seroma (fluid accumulation near a surgical incision) made them feel self-conscious.^33^ Further factors identified as barriers are detailed in Appendix F of the Supplementary File.

### Theme 3: The impact of pre- and post-operative interventions on a person’s physical and psychological health

In total, 12 papers reported that participants experienced health benefits as a result of participation in the physical activity interventions.^22–24, 26–29, 31–35^ Specifically, these benefits could be grouped according to physical health^22–24, 26–28, 31, 33^ and psychological health.^22, 23, 26, 27, 31–35^ There was variation in the rationale for physical health benefits, depending on the type of physical activity undertaken. High intensity aerobic exercise reported improved cardiovascular fitness and increased stamina as a benefit,^23, 24^ while stretching-based exercise interventions such as yoga, reported improved flexibility, reduction in tension and increased mobility.^26–28, 31^ Two papers mentioned increased energy levels,^23^ with one of these papers mentioning reduction in cancer-related fatigue, as a consequence of physical activity.^24^ This conflicts with the majority of papers, which reported fatigue as a preventor of adherence to the physical activity.

Three papers reported a reduction in pain, or an increased ability to manage pain, as a physical benefit of post-operative exercise.^27, 28, 33^ This physical benefit was reported less frequently compared to other physical health benefits. Osypiuk *et al.* reported that all of their participants noticed reduced physical tension, improved strength and flexibility, but only 60% noticed a decrease in pain.^27^ The pre-operative study by Brahmbhatt *et al.* reported that patients who had undertaken a physical activity intervention prior to surgery, recovered more quickly post-operatively; they included perspective from a breast cancer patient who was able to *“lift her arms over [her] head, one day after breast reconstruction surgery, which normally takes months after surgery to achieve”*.^22^ Another study echoed this when discussing baseline levels of exercise undertaken independently prior to diagnosis, to support patients in better tolerating surgical treatment.^34^

> “People told me ‘you seem to endure the treatment course well because you had exercised before’ and I also feel the same way”.^34^

Psychological benefits were reported by more papers than physical benefits. Reduction of stress and anxiety as a result of cancer diagnosis and treatment was a benefit of physical activity that four of the papers reported.^22–24, 35^ In particular, the worry of recurrence and upcoming treatment procedures were mentioned;

> “it caught me just in that time after diagnosis when things were pretty scary and pretty awful, and it felt like it was one of the key pieces of my plan for positivity during this whole thing, because it was setting the tone for recovery”.^22^

> *“Regaining control”* was the most discussed psychological benefit within seven of the included studies to describe the restoration of independence during the surgical pathway.^22–24, 29, 32, 34, 35^ One paper commented how the journey to regaining control had meant participants learning and accepting their new post-surgery limitations and adapting their movement accordingly; *“you have to learn to listen to the warning signals”*.^29^ The phenomenon of ‘regaining control’ directly links to the previously mentioned facilitator of participation in physical activity, motivation to recover. Breast cancer patients reported having a strong desire to recover, thus driving adherence to the prescribed physical activity.

> “It’s [exercise] going to generally make you feel better so that you see that sense of, you know, being in control of your life and I definitely feel that way”.^23^

### Theme 4: Patient recommendations for pre- and post-operative physical activity interventions

Participant recommendations for pre- and post-operative physical activity interventions were included in the results and discussion of seven papers.^22, 23, 25, 31–34^ The recommendation that physical activity interventions should be included as part of standard care when receiving surgical intervention for breast cancer was reported by participants.^23, 31, 34^ Further, in the study focusing on a pre-operative physical activity intervention, participants believed that all patients receiving surgery for breast cancer treatment should receive pre-operative exercises as part of standard care.^22^

> “I have actually said to many nurses and doctors that it [prehabilitation] should be something that’s mandatory and should be implemented at the hospital for every person going through the surgery”.^22^

A variety of recommendations were reported regarding intervention timing, design and delivery. Participants described how exercise check-ins post-surgery would be desirable, stating *“every 6 months is about right to keep you going”*.^31, 32, 34^ In addition to physical support with the exercises, increased emotional support was also recommended; one paper reported this would be more appropriate from a female exercise-lead, as participants would likely *“connect better with a female”*.^32^ This correlates with the previously reported finding that constant re-assurance was found to promote physical activity adherence.

One paper acknowledged impacts on levels of concentration, both post-operatively and whilst in receipt of chemotherapy treatment, stating *“your concentration is shot after cancer when you are having treatment”*.^36^ This participant suggested that a simple, interactive, *“less is more”* design for a remotely-delivered physical activity intervention would be more suitable, where clear answers would be given if someone had questions about the exercise.^36^ Another participant, in the same paper, discussed preferences towards content that was life-affirming and encouraging having recognised the need for *“some encouragement, because it’s such a difficult time, not just ‘do this (exercise)’ but ‘how are you finding it?’”*.^36^ In-built, personable elements to the physical activity content was viewed favourably, regardless of whether the intervention was conducted in-person, remotely or a combination of both.

## Discussion

This review aimed to further build on the limited evidence concerning the perspectives of patients undergoing surgery for breast cancer. By synthesising patient perspectives and capturing lived-experiences across the entire surgical journey, this systematic review (i) sheds new light on factors promoting and preventing engagement with physical activity and (ii) offers unique insight into the physical and psychological benefits of pre- and post-operative exercise interventions. Finally, the synthesised patient-reported recommendations could be used to inform the inclusion of physical activity within the standard care pathway of a vulnerable surgical cohort.

There was a lack of studies focused on delivering a pre-operative physical activity intervention. This differs from the existing literature base involving surgery for other types of cancers, including colorectal cancer,^37, 38^ lung cancer,^39, 40^ and oesophageal cancer.^41, 42^ When comparing the typical breast cancer pre-operative period, evidence suggests that this is typically much shorter than that of other oncological surgery types.^43^ For lung cancer patients, the British Thoracic society recommends for surgery to take place within 8 weeks of diagnosis,^44^ and for colorectal cancer a 6 week pre-operative wait is considered safe;^45^ both of these surgeries occur following longer time periods compared to the typical 30 day pre-operative period for breast cancer patients.^43^ The authors recognise that this could provide an explanation for fewer pre-operative studies for this cohort.

Despite this, one study proved that pre-operative exercise interventions had positive benefits, such as faster post-operative recovery.^22^ Future studies may seek to explore the pre-operative timeframe thus providing a stronger evidence-base for conclusions to be drawn on the feasibility of physical activity prior to breast cancer surgery. In addition to this, studies researching the entire surgical pathway, both pre- and post-operative, would provide in-depth insight. At the time of searching, the authors found no studies that researched both pre- and post-operative physical activity, indicating a gap in the current literature for breast cancer surgery.

Motivation to recover was a key factor in promoting engagement with the prescribed physical activity for patients with breast cancer during the surgical pathway. Furthermore, patients reported re-gaining control of their lives as a consequence of undertaking the physical activity. This echoes the wider literature of surgical behaviour change,^46, 47^ as well as during breast cancer treatment and recovery.^48^ A qualitative study by Drageset *et al.* found that breast cancer patients coped with their diagnosis by focusing on something else, taking things step-by-step and trying to return to normal so they could remain in control of their life.^49^ Other studies have found similar results that an adaptive coping response resulted in lower cancer distress, anxiety, depression and improved quality of life.^50, 51^ The idea that this behaviour change can then translate into intervention adherence has also been found to be true in other surgical procedures.^47, 52^

The evidence presented in this systematic review highlights the need for encouragement of health behaviour changes in patients within the surgical pathway, given that they are successful in promoting recovery. The term “teachable moment” is used by researchers to describe the proposal of lifestyle changes or health-behaviour changes to reduce risk.^53^ The utilisation of teachable moments has been shown to promote health behaviour change in a number of disciplines including smoking cessation, alcohol intake in adolescents, cancer screening and pregnancy care.^54–56^ Teachable moments have also been recognised at the point of cancer diagnosis as a useful tool for encouraging behaviour change.^56^ However, evidence suggests that these teachable moments are not being delivered to breast cancer patients due to lack of physician time, uncertainty on the appropriate health-behaviour message or lack of training in health behaviour counselling; resulting in breast cancer patients not meeting the desired lifestyle behaviours.^57–59^

Despite the paucity of qualitative evidence available on the subject, the studies included in this review reported results from an appropriate sample size, reflective of a population of patients with breast cancer across the globe. However, the authors acknowledge that there were some limitations in this systematic review. While all studies included patient characteristics to state the type of breast cancer surgery and whether patients were pre-or post-operative in their surgical journey, most studies did not have indication of which study participants said which quotations; this additional layer of understanding therefore lacked trends that could be reported specific to surgery type, time-post surgery, age, sex, or ethnicity, as this data was not available. Evidence based co-design studies may further inform successful implementation of physical activity into standard care for surgical breast cancer patients.

## Conclusion

This systematic review collated qualitative patient opinions from 14 studies, on the impact, delivery and implementation of physical activity interventions for breast cancer patients before and after surgery. Data analysis found that physical activity yielded both physical and psychological benefits for patients undergoing breast cancer surgery. The reported facilitators of patient participation in physical activity, such as motivation, fear of recurrence and group exercise, should be utilised and transferred into teachable moments within the surgical pathway to promote health behaviour change and improve surgical recovery. Evidence based co-design studies may further inform successful implementation of physical activity into standard care for surgical breast cancer patients.

## Declarations

### Funding

This work was undertaken as part of a Masters of Pharmacy Undergraduate degree (author LH, supervised by author AR-B); there was no funding for this work.

## Data Availability

The data that support the findings of this study are available from the corresponding author upon reasonable request.

## Notes

### Competing Interest Statement

The authors have declared no competing interest.

