## Supplementary File for "Prescribing pre- and post-operative physical activity interventions for people undergoing breast cancer surgery: a qualitative systematic review"

Howe *et al.* 2023.

**SUPPLEMENTARY FILE**

Items:

Appendix A: PRISMA 2020 checklist

Appendix B: Database search terms

Appendix C: Blank data extraction form

Appendix D: Study quality

Appendix E: Full report of factors that promoted adherence to physical activity interventions

Appendix F: Full report of factors that prevented adherence to physical activity interventions

Appendix G: Full report of physical and psychological benefits experienced as a result of physical activity intervention

Appendix H: Full report of physical and psychological benefits that patients undergoing breast cancer surgery experienced as a result of physical activity interventions

Appendix I: Full report of participant recommendations for pre- and post-operative physical activity interventions

Appendix A: PRISMA Checklist

| **Section and Topic** | **Item #** | **Checklist item** | **Location where item is reported** |
| --- | --- | --- | --- |
| **TITLE** | | |  |
| Title | 1 | Identify the report as a systematic review. |  |
| **ABSTRACT** | | |  |
| Abstract | 2 | See the PRISMA 2020 for Abstracts checklist. |  |
| **INTRODUCTION** | | |  |
| Rationale | 3 | Describe the rationale for the review in the context of existing knowledge. |  |
| Objectives | 4 | Provide an explicit statement of the objective(s) or question(s) the review addresses. |  |
| **METHODS** | | |  |
| Eligibility criteria | 5 | Specify the inclusion and exclusion criteria for the review and how studies were grouped for the syntheses. |  |
| Information sources | 6 | Specify all databases, registers, websites, organisations, reference lists and other sources searched or consulted to identify studies. Specify the date when each source was last searched or consulted. |  |
| Search strategy | 7 | Present the full search strategies for all databases, registers and websites, including any filters and limits used. |  |
| Selection process | 8 | Specify the methods used to decide whether a study met the inclusion criteria of the review, including how many reviewers screened each record and each report retrieved, whether they worked independently, and if applicable, details of automation tools used in the process. |  |
| Data collection process | 9 | Specify the methods used to collect data from reports, including how many reviewers collected data from each report, whether they worked independently, any processes for obtaining or confirming data from study investigators, and if applicable, details of automation tools used in the process. |  |
| Data items | 10a | List and define all outcomes for which data were sought. Specify whether all results that were compatible with each outcome domain in each study were sought (e.g. for all measures, time points, analyses), and if not, the methods used to decide which results to collect. | N/A |
|  | 10b | List and define all other variables for which data were sought (e.g. participant and intervention characteristics, funding sources). Describe any assumptions made about any missing or unclear information. | N/A |
| Study risk of bias assessment | 11 | Specify the methods used to assess risk of bias in the included studies, including details of the tool(s) used, how many reviewers assessed each study and whether they worked independently, and if applicable, details of automation tools used in the process. |  |
| Effect measures | 12 | Specify for each outcome the effect measure(s) (e.g. risk ratio, mean difference) used in the synthesis or presentation of results. | N/A |
| Synthesis methods | 13a | Describe the processes used to decide which studies were eligible for each synthesis (e.g. tabulating the study intervention characteristics and comparing against the planned groups for each synthesis (item #5)). | N/A |
|  | 13b | Describe any methods required to prepare the data for presentation or synthesis, such as handling of missing summary statistics, or data conversions. | N/A |
|  | 13c | Describe any methods used to tabulate or visually display results of individual studies and syntheses. |  |
|  | 13d | Describe any methods used to synthesize results and provide a rationale for the choice(s). If meta-analysis was performed, describe the model(s), method(s) to identify the presence and extent of statistical heterogeneity, and software package(s) used. | N/A |
|  | 13e | Describe any methods used to explore possible causes of heterogeneity among study results (e.g. subgroup analysis, meta-regression). | N/A |
|  | 13f | Describe any sensitivity analyses conducted to assess robustness of the synthesized results. | N/A |
| Reporting bias assessment | 14 | Describe any methods used to assess risk of bias due to missing results in a synthesis (arising from reporting biases). | N/A |
| Certainty assessment | 15 | Describe any methods used to assess certainty (or confidence) in the body of evidence for an outcome. | N/A |
| **RESULTS** | | |  |
| Study selection | 16a | Describe the results of the search and selection process, from the number of records identified in the search to the number of studies included in the review, ideally using a flow diagram. |  |
|  | 16b | Cite studies that might appear to meet the inclusion criteria, but which were excluded, and explain why they were excluded. |  |
| Study characteristics | 17 | Cite each included study and present its characteristics. |  |
| Risk of bias in studies | 18 | Present assessments of risk of bias for each included study. |  |
| Results of individual studies | 19 | For all outcomes, present, for each study: (a) summary statistics for each group (where appropriate) and (b) an effect estimate and its precision (e.g. confidence/credible interval), ideally using structured tables or plots. | N/A |
| Results of syntheses | 20a | For each synthesis, briefly summarise the characteristics and risk of bias among contributing studies. | N/A |
|  | 20b | Present results of all statistical syntheses conducted. If meta-analysis was done, present for each the summary estimate and its precision (e.g. confidence/credible interval) and measures of statistical heterogeneity. If comparing groups, describe the direction of the effect. | N/A |
|  | 20c | Present results of all investigations of possible causes of heterogeneity among study results. | N/A |
|  | 20d | Present results of all sensitivity analyses conducted to assess the robustness of the synthesized results. | N/A |
| Reporting biases | 21 | Present assessments of risk of bias due to missing results (arising from reporting biases) for each synthesis assessed. |  |
| Certainty of evidence | 22 | Present assessments of certainty (or confidence) in the body of evidence for each outcome assessed. |  |
| **DISCUSSION** | | |  |
| Discussion | 23a | Provide a general interpretation of the results in the context of other evidence. |  |
|  | 23b | Discuss any limitations of the evidence included in the review. |  |
|  | 23c | Discuss any limitations of the review processes used. |  |
|  | 23d | Discuss implications of the results for practice, policy, and future research. |  |
| **OTHER INFORMATION** | | |  |
| Registration and protocol | 24a | Provide registration information for the review, including register name and registration number, or state that the review was not registered. |  |
|  | 24b | Indicate where the review protocol can be accessed, or state that a protocol was not prepared. |  |
|  | 24c | Describe and explain any amendments to information provided at registration or in the protocol. | N/A |
| Support | 25 | Describe sources of financial or non-financial support for the review, and the role of the funders or sponsors in the review. |  |
| Competing interests | 26 | Declare any competing interests of review authors. |  |
| Availability of data, code and other materials | 27 | Report which of the following are publicly available and where they can be found: template data collection forms; data extracted from included studies; data used for all analyses; analytic code; any other materials used in the review. | Appendices |

Appendix B: Database search terms

**Full search strategy for all journals = [all terms in column 1 linked using ‘OR] AND [all terms in column 2 linked using ‘OR’] AND [all terms in column 3 linked using ‘OR’] AND [all terms in column 4 linked using ‘OR’]**

Table 4: OVID platforms search strategy (Embase and Medline)

| 1. A prescription for Physical activity | 2. To support | 3. Breast cancer elective surgery | 4. Qualitative systematic review |
| --- | --- | --- | --- |
| exp exercise/ physical activity.mp. exp physical activity/ exercise.mp. aerobic exercise.mp. exp aerobic exercise/ swim*/ run*/ walk*/ cycl*/ exp circuit based exercise/ exp endurance training/ exp resistance training/ | exp preoperative/ exp postoperative/ exp perioperative/ preoperative care.mp. postoperative care.mp. perioperative care.mp. surgical pathway.mp. | exp Mastectomy/ mastectomy.mp. exp breast cancer surgery/ lumpectomy.mp. breast cancer surgery.mp. breast cancer patient.mp. | exp qualitative research/ qualitative methodologies/ qualitative analysis.mp. focus group.mp. semi-structured interview.mp. |

Table 5: PsycINFO search strategy

| 1. A prescription for Physical activity | 2. To support | 3. Breast cancer elective surgery | 4. Qualitative systematic review |
| --- | --- | --- | --- |
| physical activity.mp. exp exercise/ exp physical activity/ exp sports/ | peri-operative.mp. post-operative.mp. pre-operative.mp. surgical pathway.mp. | exp surgery/ exp breast neoplasms/ breast cancer surgery.mp. exp Mastectomy/ mastectomy.mp. | exp Qualitative measures/ exp qualitative methods/ qualitative.mp. qualitative research/ |

Table 6: Scopus search strategy

| 1. A prescription for Physical activity | 2. To support | 3. Breast cancer elective surgery | 4. Qualitative systematic review |
| --- | --- | --- | --- |
| TITLE-ABS-KEY ( exercise OR "physical activity" OR physical activity OR sport OR swim* OR run* OR cycl* OR walk* OR "fitness" OR "exercise" OR "aerobic exercise" OR training ) | TITLE-ABS-KEY ( "peri-operative” OR "post-operative" OR “pre-operative” OR “breast cancer recovery” ) | TITLE-ABS-KEY ( “mastectomy” OR "breast cancer surgery" OR “lumpectomy” OR breast cancer surgery ) | TITLE-ABS-KEY ( qualitative OR "qualitative analysis" OR "focus group" OR interview OR "semi-structured interview” |

Table 7: CINAHL search strategy

| 1. A prescription for Physical activity | 2. To support | 3. Breast cancer elective surgery | 4. Qualitative systematic review |
| --- | --- | --- | --- |
| (MH "Resistance Training") OR (MH "Aerobic Exercises+") OR (MH "Group Exercise") OR (MH "Sport Specific Training") OR "physcial activity or exercise" OR (MH "Core Exercises") OR (MH "Open Kinetic Chain Exercises") OR (MH "Therapeutic Exercise") OR (MH "Aquatic Exercises") OR (MH "Exercise Positions") OR (MH "Upper Extremity Exercises") OR (MH "Lower Extremity Exercises") OR (MH "Exercise+") | "prevention or intervention or treatment or program" OR (MH "Program Development") OR (MH "Internet-Based Intervention"  AND  (MH "Perioperative Nursing") OR (MH "Perioperative Care") OR (MH "Postoperative Care") OR (MH "Preoperative Care") OR (MH "Enhanced Recovery After Surgery") OR (MH "Preoperative Period") | (MH "Breast Reconstruction") OR (MH "Lumpectomy") OR (MH "Carcinoma, Ductal, Breast/SU") OR (MH "Breast Neoplasms/SU") OR "mastectomy or breast surgery or breast removal" | (MH "Qualitative Studies+") OR "qualitative research or qualitative study or qualitative methods or interview" |

Appendix C: Blank data extraction form

Table 2: Study characteristics. Adapted from a data extraction form featured in a systematic review by the research team

| **Authors** | **Year of publication** | **Country of publication** | **Method of Qualitative data collection** | **Number of participants** | **Participant characteristics** | **Reported Surgery Type(s)* (number of participants)** | **Delivery of exercise intervention** | **Detail of physical activity undertaken** | **Timepoint within surgical pathway** | | **How long pre- or post- surgery** | **Any additional input alongside physical activity?** | **Control Group?** |
| --- | --- | --- | --- | --- | --- | --- | --- | --- | --- | --- | --- | --- | --- |
|  |  |  |  |  |  |  |  |  | **Pre-operative** | **Post-operative** |  |  |  |
| Key: ...... | | | | | | | | | | | | | |

Appendix D: Study quality

Table 3: Critical appraisal of the methodological quality of the included studies. Conducted using the CASP checklist.

| **Study Authors** | Q1- Was there a clear statement of the aims of the research? | Q2 - Is qualitative methodology appropriate? | Q3- Was the research design appropriate to address the aims of the research? | Q4- Was the recruitment strategy appropriate to the aims of the research? | Q5 - Was the data collected in a way that addressed the research issue? | Q6- Has the relationship between the researcher and participants been reported? | Q7- Have ethical issues been taken into consideration? | Q8- Was the data analysis sufficiently rigorous? | Q9- Is there a clear statement of findings? | Q10- How valuable is the research? | **Total score** | **Methodological quality (%)** |
| --- | --- | --- | --- | --- | --- | --- | --- | --- | --- | --- | --- | --- |
| Osypiuk K *et al.* ^33^ | 0 | 0 | 0 | 1 | 1 | 1 | 1 | 0 | 1 | 1 | **6** | **60** |
| Fu M *et al.*^37^ | 1 | 0 | 0 | 1 | 0 | 1 | 1 | 0 | 1 | 0 | **5** | **50** |
| Osypiuk K *et al.* ^40^ | 0 | 1 | 1 | 1 | 1 | 0 | 0 | 1 | 1 | 1 | **7** | **70** |
| Balneaves L *et al.^34^* | 1 | 1 | 0 | 1 | 1 | 1 | 1 | 1 | 0 | 1 | **8** | **80** |
| Enblom A *et al*.^35^ | 0 | 1 | 1 | 1 | 1 | 1 | 0 | 1 | 0 | 1 | **7** | **70** |
| Brennan L *et al.*^41^ | 1 | 1 | 1 | 1 | 1 | 0 | 1 | 1 | 1 | 1 | **9** | **90** |
| Brahmbhatt P *et al*.^42^ | 1 | 1 | 1 | 1 | 1 | 0 | 1 | 1 | 1 | 1 | **9** | **90** |
| Hubbard G *et al.*^43^ | 1 | 1 | 0 | 0 | 1 | 0 | 1 | 1 | 1 | 0 | **6** | **60** |
| Fazzino T *et al*.^31^ | 1 | 1 | 0 | 1 | 0 | 0 | 0 | 1 | 0 | 0 | **4** | **40** |
| Kim S *et al.*^39^ | 1 | 1 | 1 | 1 | 1 | 1 | 0 | 1 | 1 | 1 | **9** | **90** |
| Rees S *et al*.^38^ | 1 | 1 | 1 | 0 | 1 | 1 | 1 | 1 | 1 | 1 | **9** | **90** |
| Yeon S *et al.^32^* | 1 | 1 | 1 | 1 | 1 | 1 | 1 | 1 | 1 | 1 | **10** | **100** |
| Ray H *et al.*^36^ | 0 | 1 | 1 | 0 | 1 | 1 | 1 | 1 | 1 | 0 | **7** | **70** |
| Larsson I *et al*.^44^ | 0 | 1 | 1 | 1 | 1 | 0 | 0 | 1 | 1 | 1 | **7** | **70** |
| **Total for question** | **9** | **12** | **9** | **11** | **12** | **8** | **9** | **12** | **11** | **10** | **Average = 7.35** | **Average = 73.5%** |

Appendix E: Full report of factors that promoted adherence to physical activity interventions

Table 8: Full identification of all factors which promoted physical activity intervention adherence for breast cancer surgical patients, as reported in the 14 included qualitative studies.

| **Paper** | **Factors which promoted adherence** |
| --- | --- |
| Osypiuk *et al.* (2019) | - Easy exercises - Integration into daily life |
| Fu M *et al.* (2021) | - Easy exercises - Immediate feedback - independence |
| Osypiuk K *et al. (*2020) | - group work - “less is more” approach |
| Balneaves L *et al.* (2014) | - Family support - Fear of cancer returning and wanting to do everything they can - Group-based setting - Motivation/support from trainers - Wanting to return to normal - Different from other programmes as not focused on weight loss |
| Enblom A *et al.* (2017) | - Others could not see movements under water - More painless to exercise in water - Social interaction - Access to private dressing rooms |
| Brennan L *et al.* (2020) | - Motivation and desire to recover - Peer support from other breast cancer patients - “less is more” approach to information given as concentration is poor post-cancer treatment - Knowing the intervention came from the hospital |
| Brahmbhatt P *et al.* (2020) | - Easy to follow - Individualisation according to fitness level - Being able to travel and take exercises with them (home-based option) - Receiving feedback from in-person instruction - Weekly phone conversations and exercise logs to track progress - Oncology trained PTs |
| Hubbard G *et al.* (2018) | - Socialisation - Being around people who do not have cancer - Flexibility - Being indoors so not weather dependent - Family and friends - Weekly check up by physical activity specialist was a source of motivation |
| Fazzino T *et al.* (2016) | - Feeling accountable to group and themselves - Self-monitoring - Internal motivation - Group work |
| Kim S *et al.* (2019) | - Being “sick” made them feel like they should exercise - Not wanting to gain weight as this can cause recurrence - Interaction with other breast cancer patients - Group support - Breast cancer specific exercises |
| Rees S *et al.* (2020) | - Re-assurance - Making progress and graduating to harder exercises - Choice of exercise - Taking control of their own recovery - Support from physiotherapists |
| Yeon S *et al.* (2021) | - Accurate info on exercise according to time after surgery (tailored) - Encouragement and social support (from medical team, friends, family, fellow patients) - First-hand experience of the benefits of exercise - Expectations that exercise would reduce pain, speed up recovery, prevent relapse and manage health. |
| Ray H *et al.* (2013) | - Group/team support network - Interaction with nature |
| Larsson I *et al.* (2008) | - A wish/desire to stay normal - Getting control of the situation - Fear of negative side effects developing if they do not exercise |

Appendix F: Full report of factors that prevented adherence to physical activity interventions

Table 9: The preventors of adherence to physical activity interventions as stated in each of the 14 papers included in the narrative synthesis.

| **Paper** | **Factors which prevented adherence** |
| --- | --- |
| Osypiuk *et al.* (2019) | - Pain during exercise - Inconvenient timing of classes - Long or difficult commute - Other commitments (family/work) |
| Fu M *et al.* (2021) | - Took a lot of mental effort to learn the exercises |
| Osypiuk K *et al. (*2020) | - Pain during exercise |
| Balneaves L *et al.* (2014) | - Fatigue - Family commitments - Work commitments - “chemo brain” impaired mental concentration - Negative reaction from family and friends - fear of family members as they confused the weight loss for the progression of cancer |
| Enblom A *et al.* (2017) | None reported by authors |
| Brennan L *et al.* (2020) | - Upper limb dysfunction - Uncertainty on what exercises they can and cannot do - Treatment side effects (weakness, fatigue, sore skin) - Time and scheduling conflicts - Fear of movement - Financial barrier – could not afford to pay for private physiotherapy appointments - Technology: old age barrier to its use - Technology: overwhelmed by volume of content |
| Brahmbhatt P *et al.* (2020) | - Motivation - The weather - Lack of time (medical appointments in pre-operative period) |
| Hubbard G *et al.* (2018) | - Long travel distance - Self-conscious about their appearance following mastectomy so did not want to exercise with general public - Adjuvant treatment side effects - Family commitments - Reasons for not participating in study also mentioned: health issues, not being interested, other commitments (family and work) – these could be seen as barriers to physical activity. |
| Fazzino T *et al.* (2016) | - Did not like when group meetings (some people spoke for too long or off topic) - Participants became tired of intervention components that required substantial effort beyond what they were doing before (daily monitoring, daily physical activity) - Scheduling conflicts for weekly video calls - Internal motivation - Fatigue - Lack of sidewalks (rural study) - Extreme seasonal temperatures - Lack of exercise facilities where they live |
| Kim S *et al.* (2019) | - Cancer related fatigue (CRF) - Myths and fears surrounding cancer (cannot use arm, will develop lymphoedema, exercise can cause recurrence) - Treatment related side effects - Self-image (self-conscious due to change in appearance, uncomfortable to wear a wig to exercise in) - Relationship issues - Inadequate information (not knowing what to do or how much) - Physical limitations making them feel like an outsider compared to ‘normal people’ who do not feel the same exhaustion due to cancer |
| Rees S *et al.* (2020) | - Physical limitations (tightness in chest, sore arm, swelling) - Aches and pains - Fatigue |
| Yeon S *et al.* (2021) | - Physical constraints (limited mobility of arm, lymphoedema, feeling weak, seroma) - Psychological resistance (psychological withdrawal, exercise not prioritised) - Concerns regarding side effects of exercise - Concerns regarding injuries - Own theories regarding aetiology and symptoms of cancer related to exercise - Pain - Drain attached to body |
| Ray H *et al.* (2013) | - Personality conflict between group members (some wanted it to be more competitive than others) |
| Larsson I *et al.* (2008) | - Fatigue - Chemotherapy side effects (vomiting) - Limitations difficult to navigate as everyone is different - Pain in exercising |

Appendix G: Full report of physical and psychological benefits breast cancer patients experienced as a result of physical activity intervention.

Table 10: The participant reported physical and psychological benefits across all 14 papers

| **Paper** | **Physical benefits** | **Psychological benefits** |
| --- | --- | --- |
| Osypiuk *et al.* (2019) | - Reduction in tension - Improved strength - Improved flexibility - decreased pain (less reported than other physical measures) - increased mobility | - more relaxed - calm - peaceful |
| Fu M *et al.* (2021) | - stretches out tightness - relieves pain | None reported by authors |
| Osypiuk K *et al. (*2020) | - stretches out tightness - reduction in tension | - restoration of mind-body disconnect - calming - restores trust in their body |
| Balneaves L *et al.* (2014) | - increased strength and stamina - better energy levels - improved physical fitness | - regain inner strength - regain independence that had been lost with diagnosis - more in control - less anxious about cancer |
| Enblom A *et al.* (2017) | None reported by authors | None reported by authors |
| Brennan L *et al.* (2020) | - improved posture | - improved confidence - feeling calmer - making friends |
| Brahmbhatt P *et al.* (2020) | - earlier recovery | - provided momentum to make health behaviour they had been intending to make, in the preoperative period and the post-operative period - regained a sense of control - provided education they could then utilise post-surgery - provided a positive distraction |
| Hubbard G *et al.* (2018) | None reported by authors | - engaging in physical activity gave back a sense of control - being active stopped them worrying about recurrence |
| Fazzino T *et al.* (2016) | None reported by authors | None reported by authors |
| Kim S *et al.* (2019) | None reported by authors | - regaining sense of control - self comfort of exercising |
| Rees S *et al.* (2020) | None reported by authors | - regained control |
| Yeon S *et al.* (2021) | - reduced pain - promoted flexibility - increased amount of exercise - increased physical activities - reduced discomfort - expedited recovery | - heightened sense of purpose - want to exercise more |
| Ray H *et al.* (2013) | - improved cardiovascular endurance - more strength in shoulders, legs and core - improved strength and stamina - improved energy levels - reduced fatigue | - more conscious of rounded and healthy lifestyle - improved emotional strength - higher self-esteem, self-confidence and greater self-acceptance - improved body-image - regaining control - buffer against stress – helps with worry and anxiety - sense of peace (when exercising outdoors in nature) |
| Larsson I *et al.* (2008) | None reported by authors | - regaining control - learning new limits |

Appendix H: Full report of physical and psychological benefits patients undergoing breast cancer surgery experienced as a result of physical activity interventions

Table 11: The participant reported physical and psychological benefits across all 14 papers

| **Paper** | **Physical benefits** | **Psychological benefits** |
| --- | --- | --- |
| Osypiuk *et al.* (2019) | - Reduction in tension - Improved strength - Improved flexibility - decreased pain (less reported than other physical measures) - increased mobility | - more relaxed - calm - peaceful |
| Fu M *et al.* (2021) | - stretches out tightness - relieves pain | None reported by authors |
| Osypiuk K *et al. (*2020) | - stretches out tightness - reduction in tension | - restoration of mind-body disconnect - calming - restores trust in their body |
| Balneaves L *et al.* (2014) | - increased strength and stamina - better energy levels - improved physical fitness | - regain inner strength - regain independence that had been lost with diagnosis - more in control - less anxious about cancer |
| Enblom A *et al.* (2017) | None reported by authors | None reported by authors |
| Brennan L *et al.* (2020) | - improved posture | - improved confidence - feeling calmer - making friends |
| Brahmbhatt P *et al.* (2020) | - earlier recovery | - provided momentum to make health behaviour they had been intending to make, in the preoperative period and the post-operative period - regained a sense of control - provided education they could then utilise post-surgery - provided a positive distraction |
| Hubbard G *et al.* (2018) | None reported by authors | - engaging in physical activity gave back a sense of control - being active stopped them worrying about recurrence |
| Fazzino T *et al.* (2016) | None reported by authors | None reported by authors |
| Kim S *et al.* (2019) | None reported by authors | - regaining sense of control - self comfort of exercising |
| Rees S *et al.* (2020) | None reported by authors | - regained control |
| Yeon S *et al.* (2021) | - reduced pain - promoted flexibility - increased amount of exercise - increased physical activities - reduced discomfort - expedited recovery | - heightened sense of purpose - want to exercise more |
| Ray H *et al.* (2013) | - improved cardiovascular endurance - more strength in shoulders, legs and core - improved strength and stamina - improved energy levels - reduced fatigue | - more conscious of rounded and healthy lifestyle - improved emotional strength - higher self-esteem, self-confidence and greater self-acceptance - improved body-image - regaining control - buffer against stress – helps with worry and anxiety - sense of peace (when exercising outdoors in nature) |
| Larsson I *et al.* (2008) | None reported by authors | - regaining control - learning new limits |

Appendix I: Full report of participant recommendations for the improvement of the physical activity interventions

Table 12: Participant recommendations for improvement to physical activity interventions across the 14 studies

| **Paper** | **Participant recommendations for improvement to physical activity intervention** |
| --- | --- |
| Osypiuk *et al.* (2019) | None reported by authors |
| Fu M *et al.* (2021) | None reported by authors |
| Osypiuk K *et al. (*2020) | None reported by authors |
| Balneaves L *et al.* (2014) | - Intervention should be offered immediately after surgery - Exercise intervention should be included as part of standard care - Family and friends should be involved in intervention - Desire for a place to continue exercising after intervention ended, even if it meant paying additional fees. - Should be pitched as educational, rather than weight loss intervention |
| Enblom A *et al.* (2017) | None reported by authors |
| Brennan L *et al.* (2020) | - More access to physiotherapy services, especially follow-up after hospital discharge - Providing patients with more information about what to expect after the surgery, regarding diet, exercise and breast prostheses - Postoperative support group - Physiotherapist-led pre-operative information session - Technology: wished the avatar had looked like them- resemble someone receiving breast cancer treatment - System should be provided from the hospital so you know it is legitimate - Less is more approach since concentration is “shot” after cancer - Encouragement and reassurance from exercise trainers |
| Brahmbhatt P *et al.* (2020) | - Prehabilitation physical activity should be made available to everyone receiving surgery |
| Hubbard G *et al.* (2018) | None reported by authors |
| Fazzino T *et al.* (2016) | - Willing to pay to continue intervention 20 to 200 dollars. |
| Kim S *et al.* (2019) | - Tailoring exercise to individual - Make it fun and engaging - Allow flexibility - Needs to be breast cancer specific exercise - Every 6 month an exercise check-in would be good - Would be great to have programmes like this offered by hospital |
| Rees S *et al.* (2020) | - Emotional support needed in addition to physical exercise support - Would connect better with a female instructor/HCP |
| Yeon S *et al.* (2021) | - Want more consultation regarding symptoms, then exercises specific to this - Want more information/education in accordance with timing of surgery |
| Ray H *et al.* (2013) | None reported by authors |
| Larsson I *et al.* (2008) | None reported by authors |
